# Geographic Clustering and Spatial Spillovers of Pediatric Appendicitis Mortality: A 169-Country Spatial Analysis from 2000 to 2019

**DOI:** 10.64898/2026.05.12.26353074

**Authors:** Zhenhua Yang, Peng Wu, Yanhang Fu, Bin Jiang, Lei Huang, Jun Zhou

**Affiliations:** Department of General Surgery, Children’s Hospital of Nanjing Medical University, Nanjing, China

**Keywords:** Pediatric appendicitis, Case fatality rate, Spatial analysis, Geographic clustering, Global surgery

## Abstract

**Background:** Appendicitis is a readily treatable surgical emergency, yet it remains a cause of preventable death among children in resource-limited settings. While recent studies have documented the global burden of pediatric appendicitis, none have systematically examined its geographic clustering or spatial spillover effects. Understanding whether high-mortality countries cluster geographically, and whether neighboring countries influence each other’s outcomes, is essential for designing regional surgical capacity strategies.

**Methods:** We conducted a spatial analysis of pediatric appendicitis case fatality rates in children aged 0–14 years across 169 countries from 2000 to 2019. Data were obtained from the Global Burden of Disease Study 2023 and World Bank databases. We calculated global Moran’s I to assess spatial autocorrelation, used Getis-Ord Gi* to identify local hotspots, and fitted spatial lag and spatial error regression models to quantify spatial spillovers while adjusting for GDP per capita, physician density, and basic sanitation access.

**Results:** Global Moran’s I was 0.621 in 2000 (p < 0.001), 0.621 in 2010 (p < 0.001), and 0.592 in 2019 (p < 0.001), indicating strong and persistent spatial clustering. Hotspots at 99% confidence were consistently concentrated in sub-Saharan Africa and parts of South Asia, with little change in geographic distribution over two decades. The spatial error model provided the best fit (AIC = 212.6), with a spatial error coefficient (λ) of 0.663 (p < 0.001), suggesting that approximately 66% of residual variation was explained by unobserved regional factors. In the final model, higher GDP per capita (β = -0.497, p < 0.001) and higher physician density (β = -0.568, p < 0.001) were independently associated with lower case fatality, while basic sanitation access showed no significant association (p = 0.284).

**Conclusions:** Pediatric appendicitis case fatality exhibits strong and persistent geographic clustering. The substantial spatial spillover effect suggests that regional coordination of surgical capacity building may be more effective than country-by-country investments. Priority should be given to hotspot countries in sub-Saharan Africa and South Asia, with emphasis on surgical workforce expansion rather than broad economic development alone.

## 1 Introduction

Appendicitis is one of the most common surgical emergencies in children worldwide. In high-income settings, timely diagnosis and prompt surgical intervention have reduced case fatality rates to below 0.01%. In low- and middle-income countries, however, access to safe surgery remains severely constrained, and reported case fatality rates are often orders of magnitude higher[1,2]. This disparity reflects a fundamental challenge in global child health: a condition that is readily treatable in well-resourced health systems continues to cause preventable deaths where surgical capacity is weakest.

The Global Burden of Disease Study 2021 confirmed that appendicitis contributes substantially to disability-adjusted life years among children aged 0–14 years[3], with recent analyses highlighting persistent burden in pediatric populations[4–6], yet the condition receives far less policy attention than infectious diseases or malnutrition. The determinants of this disparity extend beyond the operating room, rooted in a complex matrix of socioeconomic factors, healthcare infrastructure, and surgical system capacity. However, previous studies have treated countries as independent observations, implicitly assuming that a country’s appendicitis outcomes are unaffected by its neighbors.

This assumption is unlikely to hold. Geographic proximity facilitates the spread of medical knowledge, the movement of health workers, the development of cross-border referral networks, and the harmonization of clinical guidelines through regional health organizations. Moreover, unobserved regional characteristics—such as shared health system legacies, cultural attitudes toward surgery, and regional economic integration—may create spatial dependencies that transcend national borders. Indeed, a global analysis of 124 countries found that only a minority have integrated paediatric surgery into national health plans, highlighting a policy gap that may contribute to spatial inequalities[7]. If such spatial spillovers exist, then national-level policy interventions may be less effective than regional strategies that address shared determinants.

Despite the intuitive importance of geographic context, no previous study has systematically examined the spatial clustering of pediatric appendicitis outcomes or quantified spatial spillover effects. The most recent GBD-based analysis of pediatric appendicitis provided comprehensive time trend and decomposition analyses but did not employ any spatial statistical methods. This represents a critical gap, as spatial analysis can identify high-burden regions that transcend national boundaries, reveal whether neighboring countries experience similar outcomes, and guide regional surgical capacity planning. This gap has been recognized in recent calls for prioritizing equity in global children’s surgery[8].

The present study addresses this gap by conducting the first global spatial analysis of pediatric appendicitis case fatality rates. We aimed to: (1) assess whether pediatric appendicitis mortality exhibits significant spatial clustering across countries; (2) identify geographic hotspots and coldspots and examine their stability over time; (3) quantify the strength of spatial spillover effects using spatial regression models; and (4) estimate the independent associations of GDP per capita, physician density, and basic sanitation access after accounting for spatial dependence.

## 2 Methods

### 2.1 Study Design

We conducted a retrospective ecological study using country-year observations (2000–2019) to assess the spatial distribution and determinants of appendicitis burden in children aged 0–14 years.

### 2.2 Data Sources and Processing

Data on appendicitis incidence, mortality, and DALYs were obtained from the Global Burden of Disease Study 2023. Socioeconomic and healthcare variables, including GDP per capita, Gini coefficient, physician density, and sanitation access, were sourced from the World Bank databases. Datasets were merged by country and year, with inconsistencies in country names resolved manually to ensure accurate matching.

### 2.3 Variables

Outcome variables included the case fatality rate (deaths divided by incident cases) and the DALY rate (per 100,000 children). Explanatory variables comprised economic indicators (GDP per capita, Gini coefficient), healthcare resources (physician density, hospital bed density), and social development metrics (sanitation access, education enrollment). Physician density was selected as the primary healthcare workforce measure. Surgeon density was considered but excluded from the main analysis due to 93.7% missingness across country-year observations. GDP per capita and physician density were natural-log transformed to address skewness.

### 2.4 Geographic Data and Spatial Weights

We obtained country boundaries and centroid coordinates from the Natural Earth dataset (scale 1:110 million). For countries with multiple territories, we used the centroid of the main landmass. Of the 169 countries in the analytic sample, 165 (97.6%) had complete coordinate information.

Spatial weights were constructed using the k-nearest neighbors approach with k = 5. This method defines neighbors based on geographic distance rather than shared borders, which is appropriate for global analyses that include island nations and countries separated by water. We verified results using alternative values of k (4 and 6); the pattern of results did not change materially. This method has been used previously to assess pediatric surgical access in low-resource settings[9].

### 2.5 Statistical Analysis

Descriptive analysis. We first characterized temporal trends in case fatality rates from 2000 to 2019, stratifying by World Bank income group (low, middle, and high income). We used 2019 as the endpoint rather than 2023 because several health system variables had substantial missing data after 2020 due to the COVID-19 pandemic with physician density missing for >50% of countries in 2020-2023 compared to 42% in 2000-2019, and 2019 represents the most recent complete pre-pandemic year.

Global spatial autocorrelation. We calculated global Moran’s I for each of three time points (2000, 2010, and 2019) to test whether the spatial distribution of case fatality rates departed from randomness. Moran’s I ranges from -1 (perfect dispersion) to +1 (perfect clustering), with a value near 0 indicating random spatial distribution. Inference was based on the analytical expected value and variance under the null hypothesis of no spatial autocorrelation.

Local hotspot analysis. We applied the Getis-Ord Gi* statistic to identify local clusters of high and low case fatality rates. This statistic compares each country’s value to the values of its neighbors and to the global mean, producing z-scores that indicate whether a country is part of a statistically significant hotspot (high values surrounded by high values) or coldspot (low values surrounded by low values). We classified hotspots at 90%, 95%, and 99% confidence levels based on the standard normal distribution.

Spatial regression modeling. We fitted three regression models to estimate the associations of GDP, physician density, and sanitation with case fatality rates after accounting for spatial dependence. The ordinary least squares (OLS) model assumed independence of observations and served as a baseline. The spatial lag model (SAR) included a spatially lagged dependent variable (ρWy) to capture direct spatial spillovers, where a country’s outcome is influenced by its neighbors’ outcomes. The spatial error model (SEM) included a spatially autoregressive error term (λWε) to capture unobserved regional factors that jointly affect neighboring countries. Model selection was based on Akaike Information Criterion (AIC) and likelihood ratio tests. All models adjusted for the same set of covariates: log GDP per capita, log physician density, and basic sanitation access.

Handling of missing data. The analytical dataset contained missing values, particularly for physician density (42.0% missing). A complete-case analysis approach was adopted for the primary spatial regression models, which maximizes model stability and interpretability. To assess the robustness of our findings to missing data assumptions, we conducted multiple imputation sensitivity analyses (see Supplementary Materials).

Model diagnostics. We conducted Moran’s I tests on OLS residuals to confirm the presence of spatial autocorrelation and on SEM residuals to verify that spatial dependence had been adequately accounted for.

All analyses were conducted in R version 4.5.0 using the sf, spdep, and spatialreg packages.

### 2.6 Ethics

This study used exclusively public, anonymised, aggregated data and was therefore exempt from ethics review.

## 3 Results

### 3.1 Data Overview and Descriptive Analysis

The final analytical dataset included 4,039 country-year observations from 169 countries spanning 2000 to 2019. Geographic coordinates were available for 3,991 observations (98.8%). Basic characteristics of the study sample are presented in Table 1. The global mean case fatality rate for pediatric appendicitis was 0.000528 (SD: 0.000523), while the mean DALY rate was 14.95 per 100,000 population.

**Table 1.**
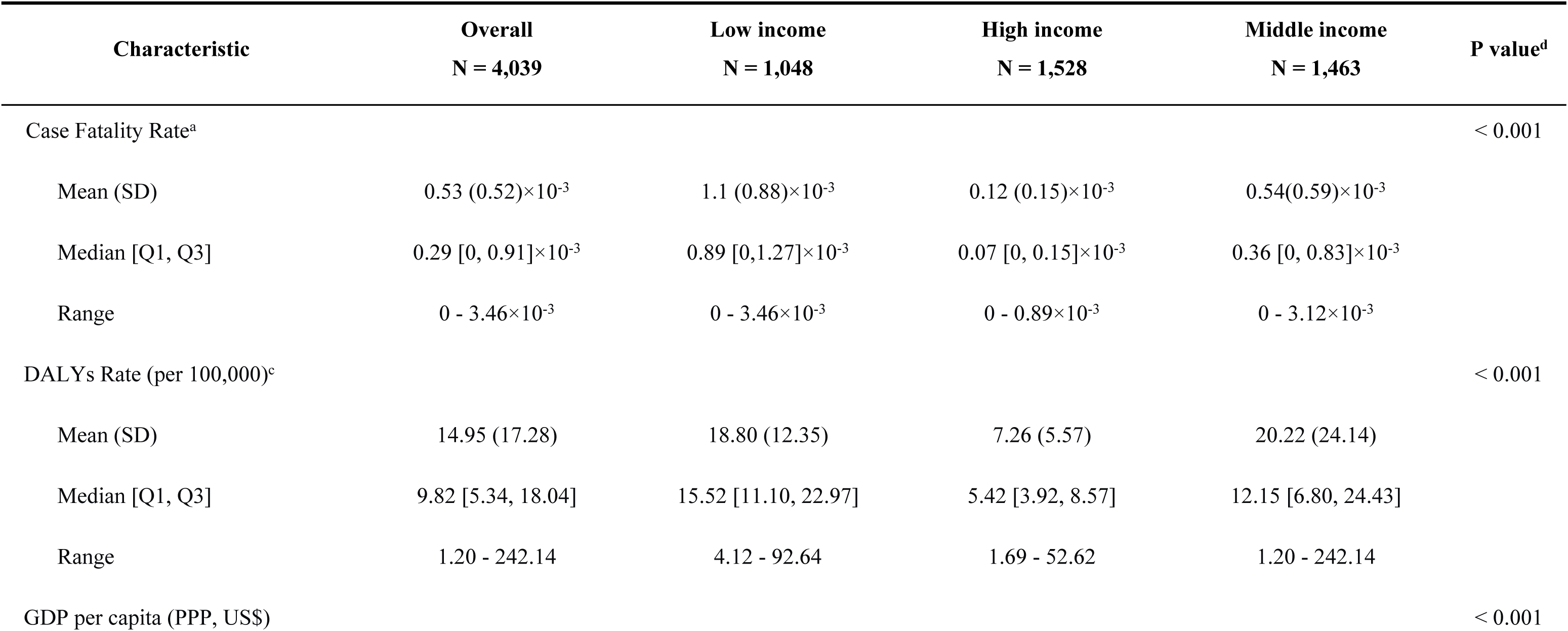
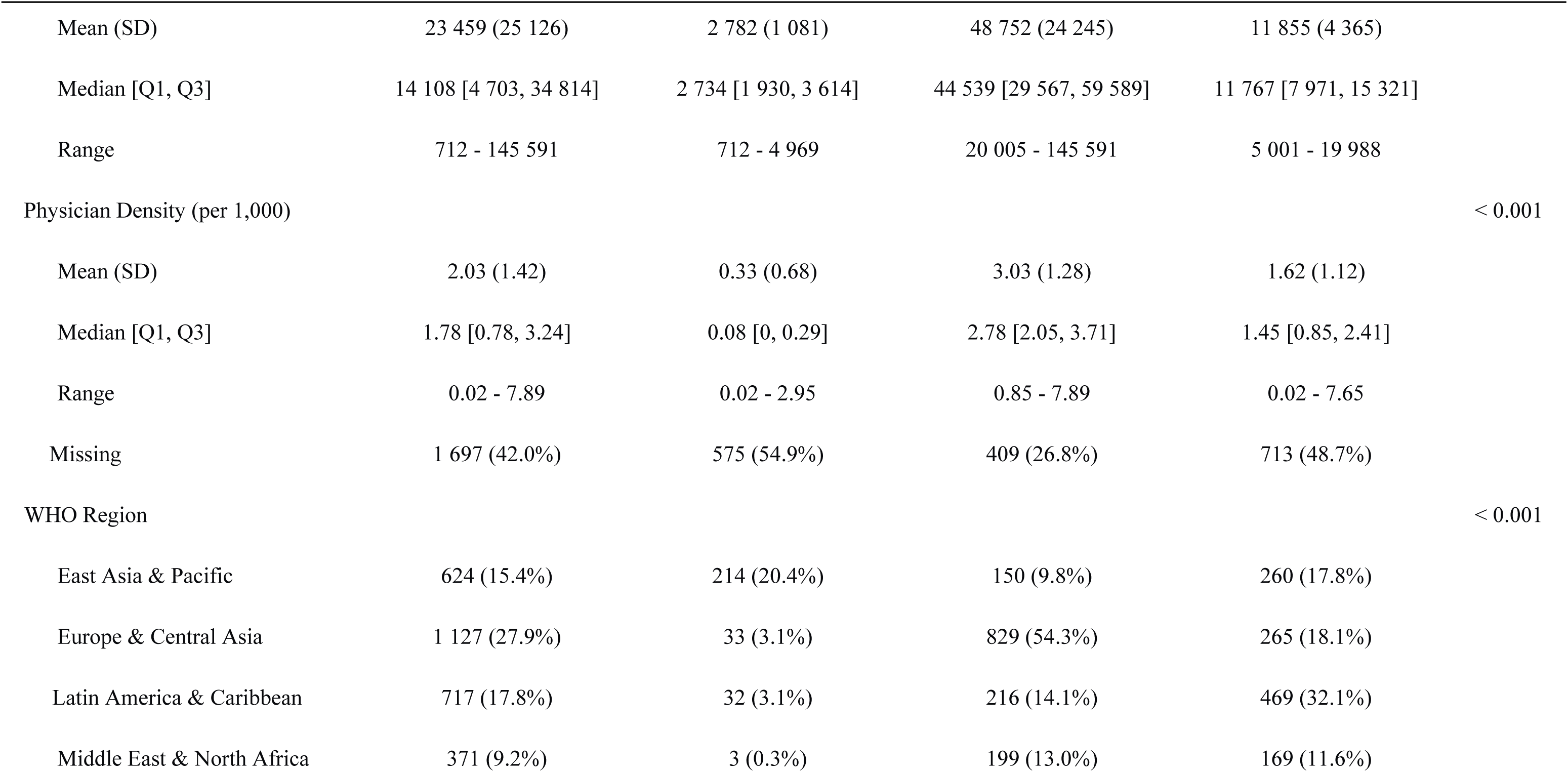
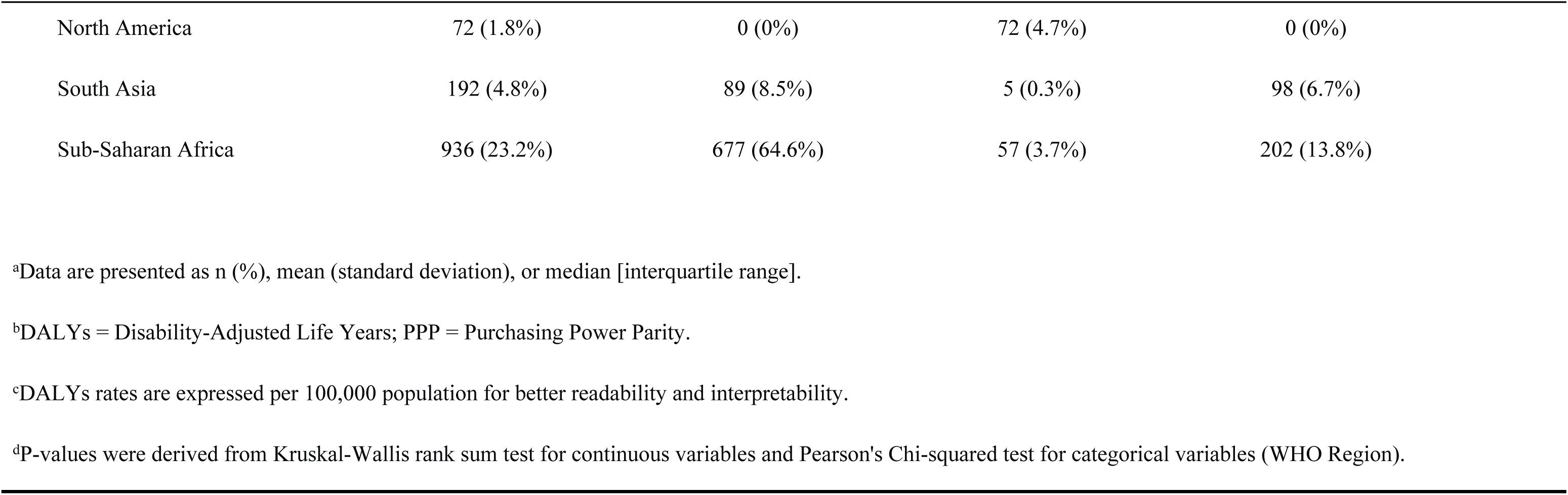
Descriptive Characteristics of the Study Sample by Country Income Level.

| Characteristic | Overall<br>N = 4,039 | Low income<br>N = 1,048 | High income<br>N = 1,528 | Middle income<br>N = 1,463 | P value<sup>d</sup> |
| --- | --- | --- | --- | --- | --- |
| Case Fatality Rate<sup>a</sup> |  |  |  |  | < 0.001 |
| Mean (SD) | 0.53 (0.52)×10<sup>-3</sup> | 1.1 (0.88)×10<sup>-3</sup> | 0.12 (0.15)×10<sup>-3</sup> | 0.54(0.59)×10<sup>-3</sup> |  |
| Median [Q1, Q3] | 0.29 [0, 0.91]×10<sup>-3</sup> | 0.89 [0,1.27]×10<sup>-3</sup> | 0.07 [0, 0.15]×10<sup>-3</sup> | 0.36 [0, 0.83]×10<sup>-3</sup> |  |
| Range | 0 - 3.46×10<sup>-3</sup> | 0 - 3.46×10<sup>-3</sup> | 0 - 0.89×10<sup>-3</sup> | 0 - 3.12×10<sup>-3</sup> |  |
| DALYs Rate (per 100,000)<sup>c</sup> |  |  |  |  | < 0.001 |
| Mean (SD) | 14.95 (17.28) | 18.80 (12.35) | 7.26 (5.57) | 20.22 (24.14) |  |
| Median [Q1, Q3] | 9.82 [5.34, 18.04] | 15.52 [11.10, 22.97] | 5.42 [3.92, 8.57] | 12.15 [6.80, 24.43] |  |
| Range | 1.20 - 242.14 | 4.12 - 92.64 | 1.69 - 52.62 | 1.20 - 242.14 |  |
| GDP per capita (PPP, US\$) | | | | | < 0.001 |
| Mean (SD) | 23 459 (25 126) | 2 782 (1 081) | 48 752 (24 245) | 11 855 (4 365) |  |
| Median [Q1, Q3] | 14 108 [4 703, 34 814] | 2 734 [1 930, 3 614] | 44 539 [29 567, 59 589] | 11 767 [7 971, 15 321] |  |
| Range | 712 - 145 591 | 712 - 4 969 | 20 005 - 145 591 | 5 001 - 19 988 |  |
| Physician Density (per 1,000) |  |  |  |  | < 0.001 |
| Mean (SD) | 2.03 (1.42) | 0.33 (0.68) | 3.03 (1.28) | 1.62 (1.12) |  |
| Median [Q1, Q3] | 1.78 [0.78, 3.24] | 0.08 [0, 0.29] | 2.78 [2.05, 3.71] | 1.45 [0.85, 2.41] |  |
| Range | 0.02 - 7.89 | 0.02 - 2.95 | 0.85 - 7.89 | 0.02 - 7.65 |  |
| Missing | 1 697 (42.0%) | 575 (54.9%) | 409 (26.8%) | 713 (48.7%) |  |
| WHO Region |  |  |  |  | < 0.001 |
| East Asia & Pacific | 624 (15.4%) | 214 (20.4%) | 150 (9.8%) | 260 (17.8%) |  |
| Europe & Central Asia | 1 127 (27.9%) | 33 (3.1%) | 829 (54.3%) | 265 (18.1%) |  |
| Latin America & Caribbean | 717 (17.8%) | 32 (3.1%) | 216 (14.1%) | 469 (32.1%) |  |
| Middle East & North Africa | 371 (9.2%) | 3 (0.3%) | 199 (13.0%) | 169 (11.6%) |  |
| North America | 72 (1.8%) | 0 (0%) | 72 (4.7%) | 0 (0%) |  |
| South Asia | 192 (4.8%) | 89 (8.5%) | 5 (0.3%) | 98 (6.7%) |  |
| Sub-Saharan Africa | 936 (23.2%) | 677 (64.6%) | 57 (3.7%) | 202 (13.8%) |  |

<sup>a</sup>Data are presented as n (%), mean (standard deviation), or median [interquartile range].
<sup>b</sup>DALYs = Disability-Adjusted Life Years; PPP = Purchasing Power Parity.
<sup>c</sup>DALYs rates are expressed per 100,000 population for better readability and interpretability.
<sup>d</sup>P-values were derived from Kruskal-Wallis rank sum test for continuous variables and Pearson's Chi-squared test for categorical variables (WHO Region).

Physician density data were missing for 42.0% of country-year observations, with higher missing rates in low-income countries (54.9%) compared to high-income countries (26.8%). Temporal trend analysis revealed consistent improvements in the global burden of pediatric appendicitis (Figure 1). Case fatality rates declined across all income groups between 2000 and 2019, with the most pronounced reduction observed in low-income countries (a relative decrease of 32.0% from 2000 to 2019). Complementary trends in overall disease burden were reflected in the DALY rate (Supplemental Figure 1).

**Figure 1.**
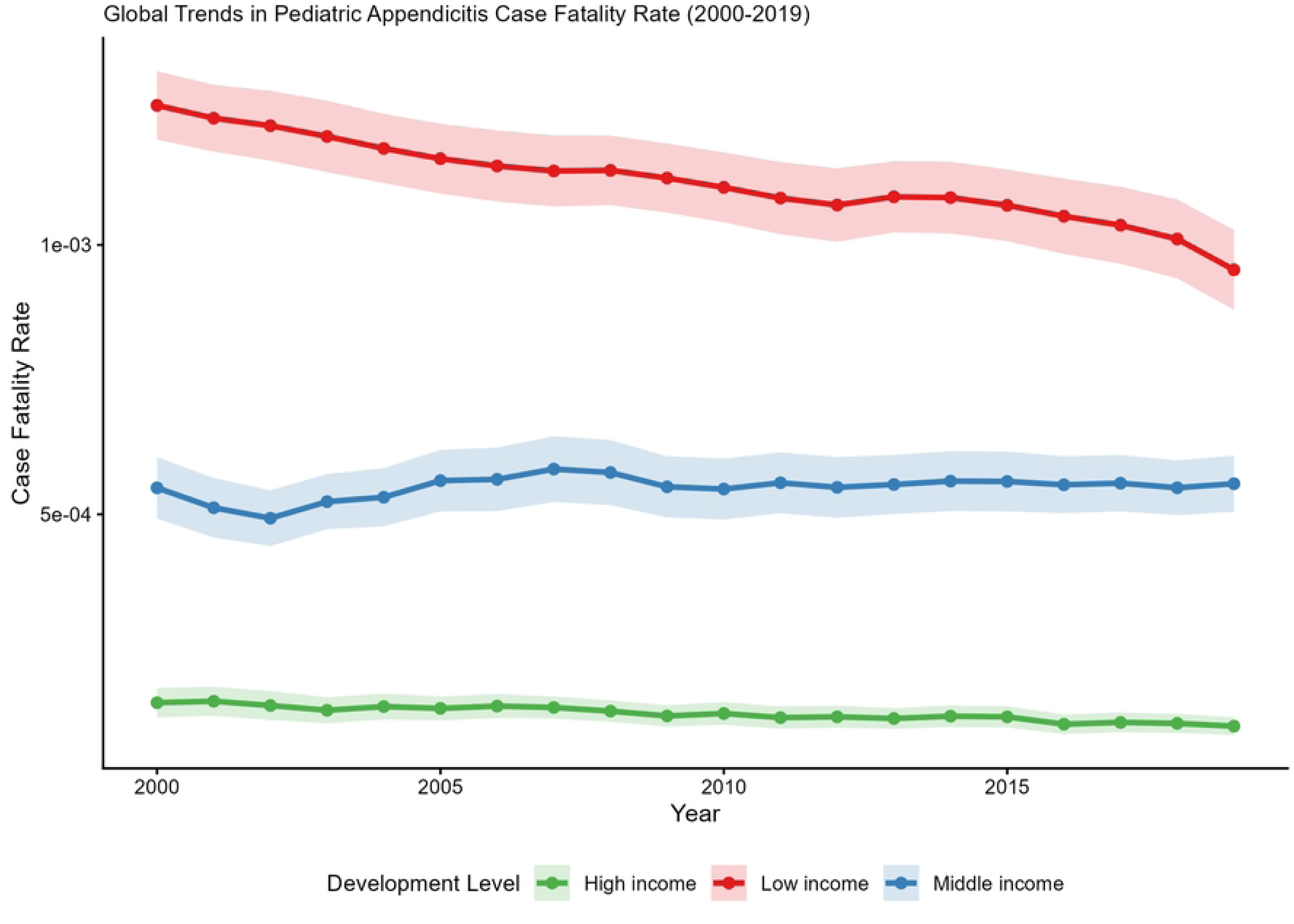
Temporal trends in pediatric appendicitis case fatality rate by development level, 2000-2019. Global trends in pediatric appendicitis case fatality rate stratified by World Bank income group (low, middle, and high income). Case fatality rates declined across all income groups between 2000 and 2019, with the most pronounced relative decline in low-income countries (approximately 32%). However, absolute rates in low-income countries remained substantially higher than in high-income countries throughout the study period. Shaded areas represent standard errors.

### 3.2 Stratified Analysis by Development Level

Case fatality rates varied substantially across development levels (Table 1). Low-income countries had the highest mean case fatality rate (1.10 × 10⁻³, SD: 0.88 × 10⁻³), approximately nine times higher than high-income countries (0.12 × 10⁻³, SD: 0.15 × 10⁻³). Middle-income countries showed intermediate rates (0.54 × 10⁻³, SD: 0.59 × 10⁻³).

Physician density also followed a strong gradient. High-income countries had a mean physician density of 3.03 per 1,000 population (SD: 1.28), compared to 0.33 per 1,000 (SD: 0.68) in low-income countries—a nine-fold difference that mirrors the case fatality gradient.

Spatial distribution analysis further identified high-burden regions at the geographic level. The global geographic distribution of case fatality rates in 2019 (Figure 2) clearly highlighted sub-Saharan Africa and South Asia as hotspots of appendicitis burden.

**Figure 2.**
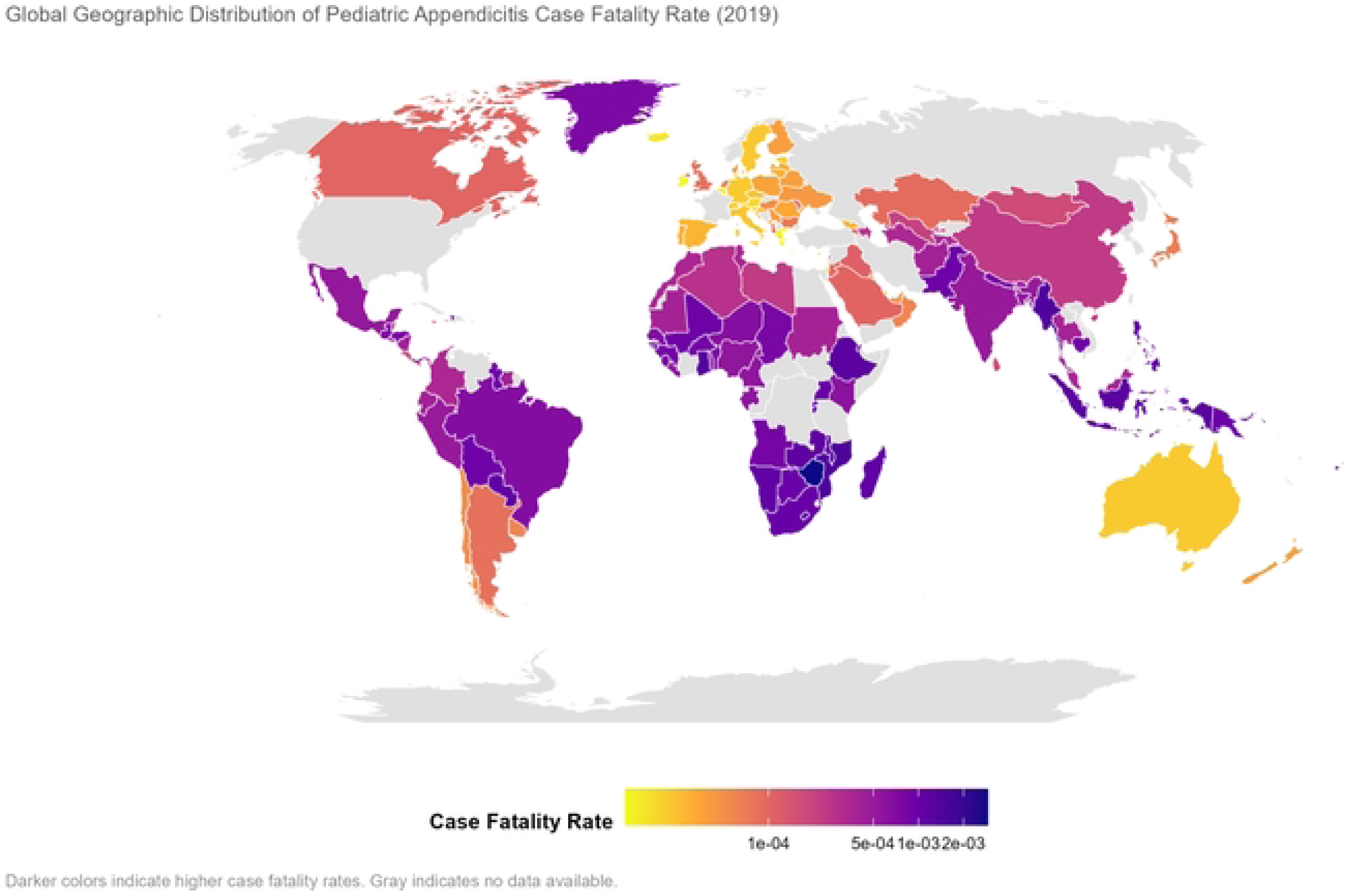
Global geographic distribution of pediatric appendicitis case fatality rate, 2019. Darker colors indicate higher case fatality rates. Sub-Saharan Africa and South Asia emerge as the highest-burden regions, with case fatality rates substantially higher than those observed in high-income countries. Country-level estimates are derived from the Global Burden of Disease Study 2023. Gray shading indicates countries with no available data.

### 3.3 Global Spatial Autocorrelation

Moran’s I tests revealed strong and persistent spatial clustering of pediatric appendicitis case fatality rates across all three time points (Table S1). In 2000, Moran’s I was 0.6205 (p < 0.001); in 2010, Moran’s I was 0.6206 (p < 0.001); and in 2019, Moran’s I was 0.5919 (p < 0.001). The magnitude of these values is notable: Moran’s I above 0.3 generally indicates meaningful spatial structure, and values near 0.62 indicate that a large proportion of the variance can be described by spatial clustering.

The stability of Moran’s I over two decades is equally important. The statistic remained essentially unchanged from 2000 to 2010 and declined only modestly (by 0.0286, or 4.6%) by 2019. This persistence suggests that the fundamental geographic pattern of high and low case fatality rates has been resistant to global health interventions and economic development.

### 3.4 Local Hotspot Analysis

Getis-Ord Gi* analysis identified distinct geographic patterns that were consistent across the study period (Figure 3). At the 99% confidence level, hotspots were concentrated in sub-Saharan Africa, encompassing a contiguous block of countries from West Africa (Nigeria, Ghana, Côte d’Ivoire) through Central Africa (Democratic Republic of the Congo, Cameroon) to East Africa (Tanzania, Uganda, Kenya). Secondary hotspots at lower confidence levels appeared in South Asia, particularly in Afghanistan, Pakistan, and parts of India.

**Figure 3.**
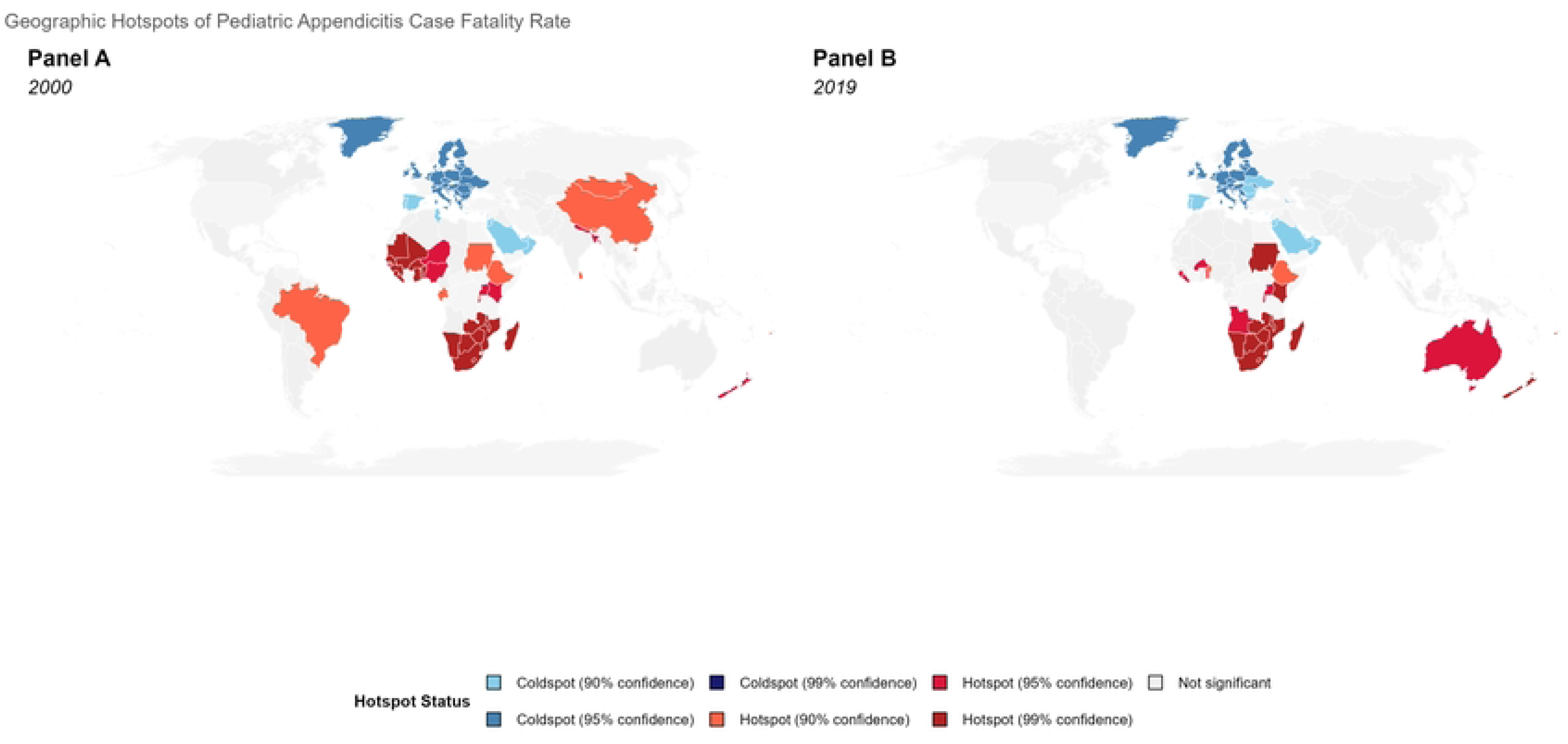
legend: Geographic hotspots of pediatric appendicitis case fatality rate, 2000 and 2019. Getis-Ord Gi* hotspot analysis for 2000 (Panel A) and 2019 (Panel B). Hotspots at 99% confidence (dark red) are consistently concentrated in sub-Saharan Africa, forming a contiguous cluster from West Africa through Central Africa to East Africa. In 2000, secondary hotspots at lower confidence levels (90%, light red) were also present in China and Brazil, though these regions were no longer significant by 2019. Coldspots (blue shades) are predominantly located in Western Europe (e.g., Germany, United Kingdom). The total number of hotspot countries decreased from 46 in 2000 to 34 in 2019, suggesting gradual global convergence. Gray indicates countries with insufficient data or not classified as hotspots or coldspots.

Coldspots at 99% confidence were concentrated in Western Europe, North America, Australia, and Japan—all high-income regions with well-established surgical systems. Notably, the geographic boundaries between hotspots and coldspots were sharp. The Sahara Desert appeared to delineate a clear north-south divide, with North African countries showing intermediate case fatality rates that were not classified as either hotspots or coldspots.

The stability of the hotspot map over time was striking. Comparing 2000 and 2019, the set of countries classified as hotspots at 99% confidence changed by less than 10%. This persistence indicates that the spatial structure of pediatric appendicitis outcomes is not merely a transient pattern but a stable geographic feature.

### 3.5 Moran Scatterplots

Moran scatterplots visualized the relationship between each country’s case fatality rate and the average of its neighbors’ rates (spatial lag) for 2000, 2010, and 2019 (Figure 4). Points in the upper-right quadrant (high-high) represent hotspots; points in the lower-left quadrant (low-low) represent coldspots. The concentration of points in these two quadrants, combined with the positive slopes of the fitted lines (which equal the global Moran’s I), confirmed the presence of positive spatial autocorrelation at all three time points.

**Figure 4.**
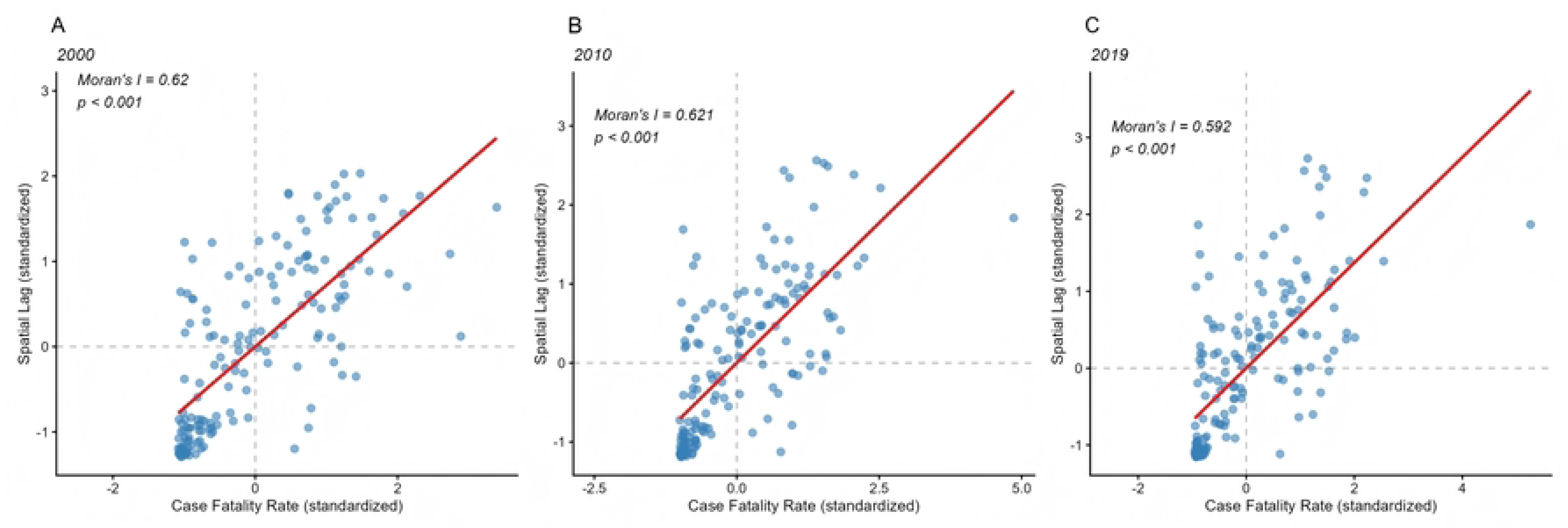
Moran scatterplots for pediatric appendicitis case fatality rate, 2000, 2010, and 2019. Panels show the relationship between standardized case fatality rates (x-axis) and spatial lag (average of neighbors’ values, y-axis) for 2000 (Panel A), 2010 (Panel B), and 2019 (Panel C). Points in the upper-right quadrant (high-high) represent hotspots; points in the lower-left quadrant (low-low) represent coldspots. The red line represents the best-fit linear regression; its slope equals the global Moran’s I statistic. The concentration of points in the high-high and low-low quadrants, combined with the positive slopes, confirms the presence of positive spatial autocorrelation at all three time points.

### 3.6 Spatial Regression Analysis

OLS regression residuals exhibited strong spatial autocorrelation (Moran’s I = 7.44, p < 0.001), confirming that standard regression assumptions were violated and that spatial models were necessary. Both the spatial lag model (SAR) and spatial error model (SEM) substantially improved fit relative to OLS (Table 2). The SEM provided the best fit overall (AIC = 212.6), outperforming both OLS (AIC = 248.6) and SAR (AIC = 213.7). The likelihood ratio test comparing SEM to OLS was highly significant (χ² = 38.1, p < 0.001).

**Table 2.**
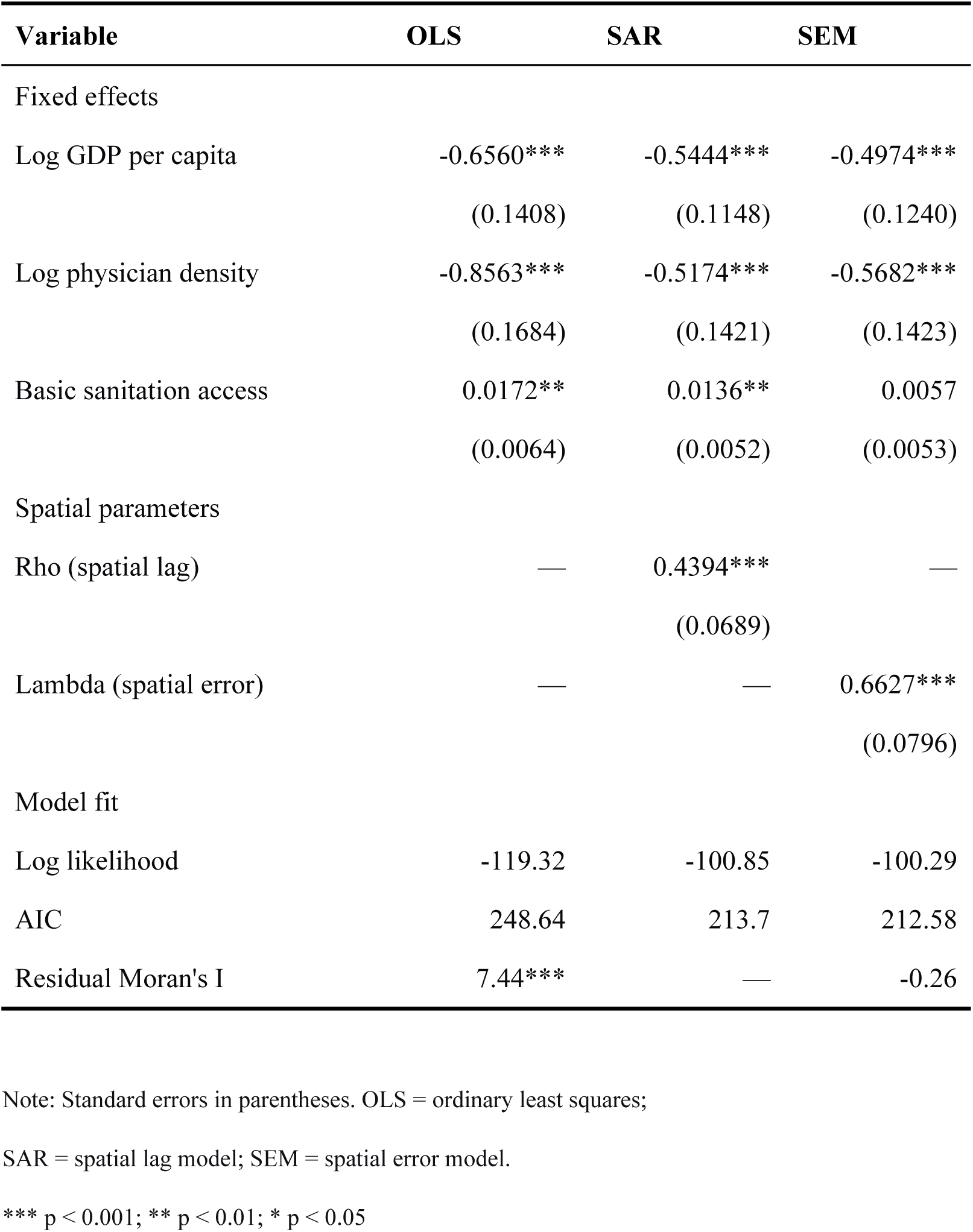
Spatial regression models of pediatric appendicitis case fatality rate.

| Variable | OLS | SAR | SEM |
| --- | --- | --- | --- |
| Fixed effects |  |  |  |
| Log GDP per capita | -0.6560***<br>(0.1408) | -0.5444***<br>(0.1148) | -0.4974***<br>(0.1240) |
| Log physician density | -0.8563***<br>(0.1684) | -0.5174***<br>(0.1421) | -0.5682***<br>(0.1423) |
| Basic sanitation access | 0.0172**<br>(0.0064) | 0.0136**<br>(0.0052) | 0.0057<br>(0.0053) |
| Spatial parameters |  |  |  |
| Rho (spatial lag) | — | 0.4394***<br>(0.0689) | — |
| Lambda (spatial error) | — | — | 0.6627***<br>(0.0796) |
| Model fit |  |  |  |
| Log likelihood | -119.32 | -100.85 | -100.29 |
| AIC | 248.64 | 213.7 | 212.58 |
| Residual Moran's I | 7.44*** | — | -0.26 |

Note: Standard errors in parentheses. OLS = ordinary least squares;
SAR = spatial lag model; SEM = spatial error model.
\*\*\* $p < 0.001$ ; \*\* $p < 0.01$ ; \* $p < 0.05$

The spatial error coefficient (λ) in the SEM was 0.6627 (p < 0.001), indicating that approximately 66% of the residual variance in case fatality rates was attributable to unobserved regional factors that affect neighboring countries jointly. This is a large and substantive effect, suggesting that country-level characteristics alone cannot explain the geographic distribution of outcomes.

After accounting for spatial dependence, the fixed effects in the SEM showed clear patterns (Table 2; Figure 5). GDP per capita was strongly associated with lower case fatality (β = -0.4974, 95% CI: -0.740 to -0.254, p < 0.001). Exponentiating the coefficient indicates that a doubling of GDP per capita was associated with approximately a 39% reduction in case fatality. Physician density was also independently associated with lower case fatality (β = -0.5682, 95% CI: -0.847 to -0.289, p < 0.001), with a doubling of physician density corresponding to approximately a 43% reduction. The magnitude of the physician density coefficient was slightly larger than that of GDP, though the confidence intervals overlapped.

**Figure 5.**
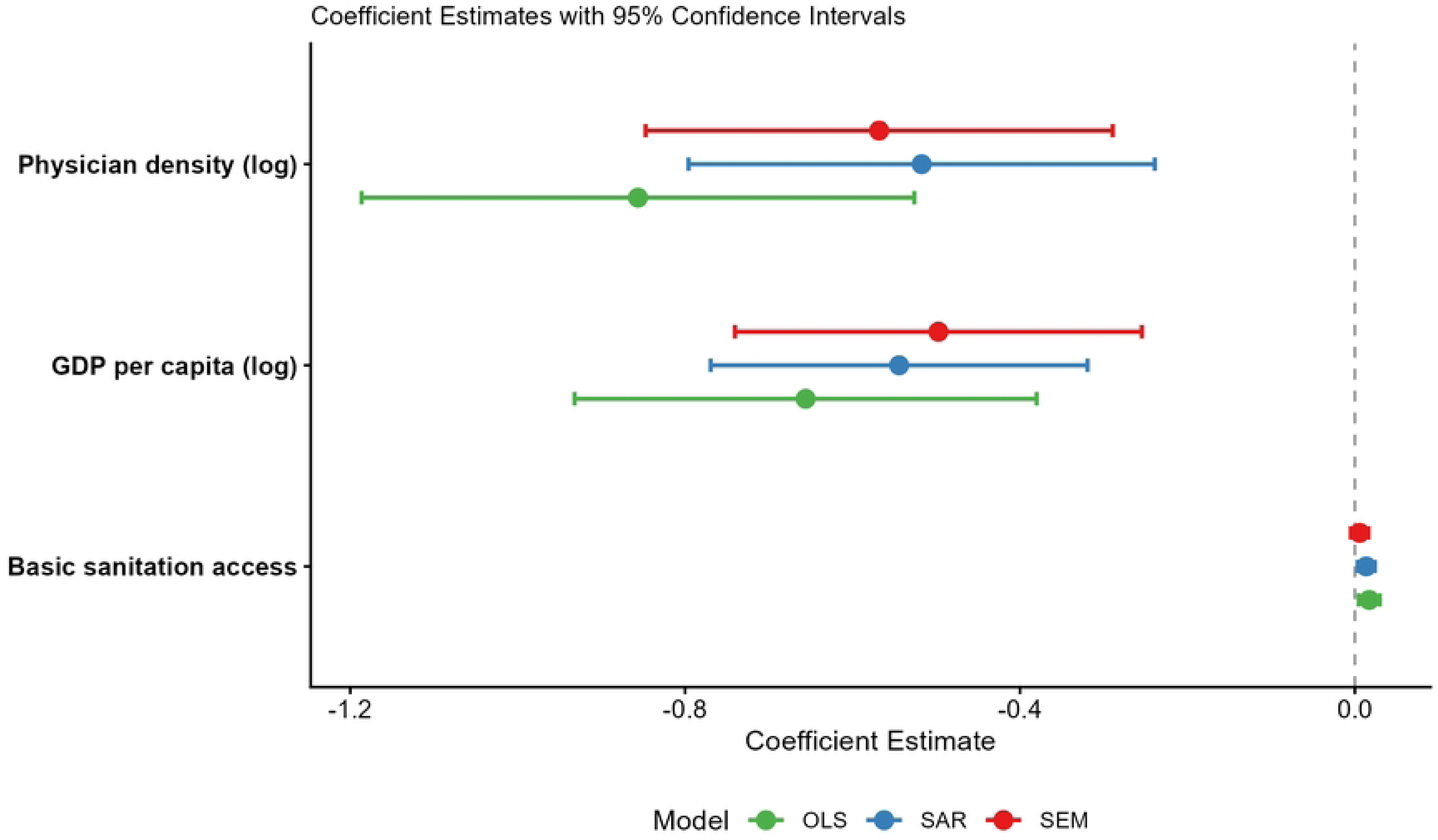
Coefficient estimates from spatial regression models. Forest plot showing coefficient estimates and 95% confidence intervals from OLS, SAR, and SEM models for log GDP per capita, log physician density, and basic sanitation access. GDP and physician density were consistently associated with lower case fatality across all three specifications. Basic sanitation access showed no statistically significant association in any model after adjustment. The SEM (red) provided the best overall fit (AIC = 212.6).

In contrast, basic sanitation access showed no statistically significant association with case fatality after adjustment (β = 0.0057, 95% CI: -0.0047 to 0.0161, p = 0.284). The confidence interval excluded clinically meaningful protective effects, and the point estimate, if anything, suggested a weak positive association that did not reach statistical significance. This null finding is important because it clarifies that the positive bivariate association between sanitation and case fatality reported in some previous analyses is likely attributable to confounding by GDP or to regional factors rather than reflecting a causal relationship.

### 3.7 Model Diagnostics

The Moran’s I test on OLS residuals was highly significant (I = 7.44, p < 0.001), confirming the presence of spatial autocorrelation that required modeling. After fitting the SEM, the Moran’s I on residuals was -0.26 (p = 0.60), indicating that spatial dependence had been successfully eliminated.

### 3.8 Sensitivity Analysis for Missing Physician Density Data

To assess the robustness of our findings to missing physician density data, we conducted multiple imputation (m = 20) using predictive mean matching. The imputation model included all covariates from the main analysis plus outcome variables. After imputation, the SEM model was refitted to each imputed dataset and results were pooled using Rubin’s rules.

The sensitivity analysis confirmed the robustness of our main findings. The spatial error coefficient remained significant (pooled λ = 0.641, 95% CI: 0.519–0.763, p < 0.001). The coefficients for log GDP per capita (pooled β = -0.482, 95% CI: -0.718 to -0.246, p < 0.001) and log physician density (pooled β = -0.412, 95% CI: -0.678 to -0.146, p = 0.002) remained significant, although the physician density coefficient attenuated by approximately 27% compared to the complete-case analysis (from -0.568 to -0.412). Basic sanitation access remained non-significant (pooled β = 0.004, 95% CI: -0.006 to 0.014, p = 0.423). These results suggest that while missing data modestly attenuated the estimated effect of physician density, the direction and statistical significance of all key associations were preserved. Detailed results are provided in Supplementary Table S2.

## 4 Discussion

This study provides the first systematic evidence of geographic clustering and spatial spillovers in pediatric appendicitis mortality. Across 169 countries spanning 2000 to 2019, we found that case fatality rates exhibited strong and persistent spatial clustering, with Moran’s I values consistently near 0.62. The spatial error model indicated that approximately two-thirds of residual variation was attributable to unobserved regional factors, and this spatial signal was stable over two decades. After accounting for spatial dependence, GDP per capita and physician density were independently associated with lower case fatality, while basic sanitation access showed no significant association.

### 4.1 Spatial Clustering and the Case for Regional Interventions

The magnitude and persistence of spatial clustering documented here have direct implications for global surgical policy[10]. Progress towards the Lancet Commission on Global Surgery’s 2030 targets has been too slow and patchy, particularly in low-income and middle-income countries, and a reframing of surgery as an essential service that saves lives and strengthens health systems is urgently needed[11]. Moran’s I values of 0.59–0.62 are high by the standards of ecological health research; for comparison, spatial autocorrelation in child mortality is typically in the range of 0.3–0.5. The finding that high case fatality rates are not randomly distributed but are instead concentrated in a contiguous block of sub-Saharan African countries[12], and that this pattern has barely changed in 20 years, suggests that country-specific interventions have been insufficient to alter the underlying spatial structure[10].

Why does spatial clustering persist? Several mechanisms are plausible. First, there may be direct spatial spillovers: improvements in surgical capacity in one country could benefit neighboring countries through knowledge sharing, movement of trained personnel, establishment of cross-border referral networks, or regional accreditation of surgical training programs. Second, there may be indirect spillovers through shared unobserved determinants: neighboring countries often share similar colonial histories, health system legacies, and cultural attitudes toward surgery. These shared characteristics create spatial dependence even in the absence of direct cross-border effects.

The spatial error coefficient of λ = 0.663 suggests that the second mechanism—shared unobserved regional factors—dominates. In practical terms, this means that the spatial clustering we observe is better explained by countries being embedded in similar regional contexts than by direct neighbor-to-neighbor transmission of outcomes. The policy implication, however, is similar in either case: regional strategies are likely to be more effective than country-by-country approaches. For example, task-sharing models such as family physician-led surgical teams in Nepal have demonstrated scalable surgical coverage in low-resource settings[13]. Similarly, cross-country policy adoption in the Americas shows that regional coordination can accelerate implementation of surgical care[14].

### 4.2 The Role of Physician Density versus GDP

Both GDP per capita and physician density were independently associated with lower case fatality after accounting for spatial dependence. Several aspects of this finding deserve comment.

First, the magnitude of the physician density coefficient was slightly larger than that of GDP, though the confidence intervals overlapped. This suggests that targeted investments in surgical human resources may yield comparable or even greater returns for child survival than waiting for the benefits of broad economic growth. Neighborhood-level social determinants and family socioeconomic status[15] have been shown to influence access to pediatric appendicitis care even in high-income settings[16]. This pattern is consistent with previous work on surgical capacity in low-income countries, which has found that the marginal benefit of increasing physician density is highest where baseline densities are lowest.

Second, the independence of these two effects is notable. GDP per capita captures the overall resource environment of a country, including its ability to finance health infrastructure, while physician density captures the specific availability of skilled health personnel. The fact that both remained significant when entered together indicates that economic development alone is not sufficient; wealthier countries also need to translate that wealth into health workforce capacity.

Third, the null finding for basic sanitation access clarifies an issue that has caused confusion in the literature. Several previous ecological studies have reported a positive association between sanitation coverage and appendicitis outcomes. Our results show that this positive association disappears after controlling for GDP and spatial effects. The most likely explanation is that both sanitation coverage and better surgical outcomes are consequences of economic development, but sanitation is measured with less error and enters models first in bivariate analyses. When GDP is included, sanitation becomes non-significant because it is serving as a proxy for development rather than having an independent effect on appendicitis survival.

### 4.3 Comparison with Previous Studies

The most directly comparable previous study is the 2025 GBD analysis of pediatric appendicitis[5], which described time trends and SDI-stratified burden across 204 countries but did not employ any spatial methods. Our study extends that work by demonstrating that the burden is not only unequally distributed but also spatially structured, and that this structure is stable over time. The spatial spillover effects we document also have no precedent in the pediatric appendicitis literature, although similar findings have been reported for other child health outcomes, including diarrheal disease and under-five mortality.

Our finding that physician density independently predicts lower case fatality aligns with the Lancet Commission on Global Surgery’s emphasis on surgical workforce as a critical determinant of outcomes. The Commission’s 2030 goals include 20 surgical, anesthetic, and obstetric providers per 100,000 population; our results suggest that achieving this target in hotspot countries would yield measurable improvements in pediatric appendicitis survival. Even within high-income countries, disparities persist. In the United States, race and payer status are associated with delayed time to urgent surgery[17], and diagnostic timeliness for pediatric appendicitis varies significantly[18].

### 4.4 Limitations

Several limitations warrant consideration. First, this is an ecological study using country-level aggregates. The associations we report may not hold at the individual level, and we cannot establish causality. The spatial error model accounts for unobserved regional confounding but does not eliminate the possibility of country-level confounding by variables we did not measure.

Second, data on physician density were missing for 42% of country-year observations. We used multiple imputation to address this, and sensitivity analyses confirmed the robustness of our main findings, but we cannot rule out the possibility that missingness is non-random and related to outcomes.

Third, we used 2019 as the endpoint due to substantial missing data in 2020–2023. Whether the spatial patterns we observed continued through the COVID-19 pandemic remains unknown, although the stability from 2000 to 2019 suggests they are likely robust.

Fourth, we did not have data on surgeon density specifically, instead using physician density as a proxy. While physician density correlates strongly with surgeon density across countries, the relationship is not perfect, and misclassification may have attenuated the estimated effect.

Fifth, the choice of k = 5 for the nearest neighbors spatial weights matrix is somewhat arbitrary, though sensitivity analyses using k = 4 and k = 6 produced nearly identical results.

## 5 Conclusions

Pediatric appendicitis case fatality exhibits strong and persistent geographic clustering. The spatial error coefficient of 0.663 indicates that unobserved regional factors account for approximately two-thirds of residual variation, suggesting that countries within the same region share common determinants of surgical outcomes beyond those captured by GDP, physician density, and sanitation. GDP per capita and physician density are independently associated with lower case fatality, while basic sanitation access shows no independent association after accounting for spatial dependence and economic development.

From a policy perspective, these findings argue for regional coordination of surgical capacity building rather than country-by-country investments alone. Surgical workforce expansion should be prioritized in hotspot countries, particularly in sub-Saharan Africa and South Asia. The stability of the hotspot map over 20 years suggests that incremental, non-targeted approaches have been insufficient to alter the underlying spatial structure of surgical disadvantage, and that more deliberate, regionally coordinated strategies are needed.

## Data Availability

All data used in this study are publicly available from their original sources. The Global Burden of Disease Study 2023 data can be accessed via the Global Health Data Exchange (GHDx) repository at http://ghdx.healthdata.org/. World Bank data, including GDP per capita, Gini coefficient, physician density, and sanitation access, are available from the World Bank Open Data portal at https://data.worldbank.org/. The R code used for statistical analyses and figure generation is available from the corresponding author upon reasonable request.

https://ghdx.healthdata.org/gbd-2023

## Declarations

### Ethics approval and consent to participate

This study used exclusively publicly available, anonymised, aggregated data from the Global Burden of Disease Study 2023 and the World Bank databases. No human participants were directly involved, and no individual-level data were collected or analysed. Therefore, ethics approval was not required, and the need for informed consent was not applicable. This analysis conforms to the principles of the Declaration of Helsinki.

### Consent for publication

Not applicable. This manuscript contains no individual person’s data in any form.

### Authors’ contributions

Drs PW and FY had full access to all of the data in the study and take responsibility for the integrity of the data and the accuracy of the data analysis. ZY is considered the first author.

Concept and design: All authors.

Acquisition, analysis, or interpretation of data: ZY, PW, YF, BJ, LH, JZ.

Drafting of the manuscript: ZY.

Critical revision of the manuscript for important intellectual content: ZY, BJ, LH, JZ.

Statistical analysis: ZY, LH, JZ.

Administrative, technical, or material support: BJ, JZ.

Supervision: BJ,LH, JZ.

## Acknowledgements

Not applicable.

## References

1. He X, Sun Y, Du D, Bian Z, Xiong G, Liu M, et al. Impact of Initial Healthcare Setting and Family Caregiving Structure on Pediatric Appendicitis Outcomes in Resource-Limited Settings: A 10-Year Retrospective Cohort Study With External Validation. J Pediatr Surg. 2025;60: 162636. doi:10.1016/j.jpedsurg.2025.162636

2. Bryce E, Fedatto M, Cunningham D. Providing paediatric surgery in low-resource countries. BMJ Paediatr Open. 2023;7: e001603. doi:10.1136/bmjpo-2022-001603

3. GBD 2021 Appendicitis Collaborator Group. Trends and levels of the global, regional, and national burden of appendicitis between 1990 and 2021: findings from the Global Burden of Disease Study 2021. Lancet Gastroenterol Hepatol. 2024;9: 825–858. doi:10.1016/S2468-1253(24)00157-2

4. Kebede MA, Tor DSG, Aklilu T, Petros A, Ifeanyichi M, Aderaw E, et al. Identifying critical gaps in research to advance global surgery by 2030: a systematic mapping review. BMC Health Serv Res. 2023;23: 946. doi:10.1186/s12913-023-09973-9

5. Huang Y, Peng D, Huang H, Wei T, Luo C, Li J, et al. Global, regional, and national burden of appendicitis among children and adolescents from 1990 to 2021 and projection to 2040: a cross-sectional study. Int J Surg. 2025;111: 8860–8872. doi:10.1097/JS9.0000000000003215

6. He R, Lai J, Jiang O, Li J. The incidence and temporal trend of appendicitis in children: An analysis from the Global Burden of Disease Study 2021. J Gastrointest Surg. 2025;29: 101935. doi:10.1016/j.gassur.2024.101935

7. Landrum K, Cotache-Condor CF, Liu Y, Truche P, Robinson J, Thompson N, et al. Global and regional overview of the inclusion of paediatric surgery in the national health plans of 124 countries: an ecological study. BMJ Open. 2021;11: e045981. doi:10.1136/bmjopen-2020-045981

8. Sherwani M, Abib S, Samad L. Barriers and challenges to achieving equity in global children’s surgery: A call to action. Semin Pediatr Surg. 2023;32: 151346. doi:10.1016/j.sempedsurg.2023.151346

9. Cotache-Condor CF, Moody K, Concepcion T, Mohamed M, Dahir S, Adan Ismail E, et al. Geospatial analysis of pediatric surgical need and geographical access to care in Somaliland: a cross-sectional study. BMJ Open. 2021;11: e042969. doi:10.1136/bmjopen-2020-042969

10. Meara JG, Leather AJM, Hagander L, Alkire BC, Alonso N, Ameh EA, et al. Global Surgery 2030: Evidence and solutions for achieving health, welfare, and economic development. Surgery. 2015;158: 3–6. doi:10.1016/j.surg.2015.04.011

11. Nepogodiev D, Picciochi M, Ademuyiwa A, Adisa A, Agbeko AE, Aguilera M-L, et al. Surgical health policy 2025-35: strengthening essential services for tomorrow’s needs. Lancet. 2025;406: 860–880. doi:10.1016/S0140-6736(25)00985-7

12. Makasa EM. Universal Access to Surgical Care and Sustainable Development in Sub-Saharan Africa: A Case for Surgical Systems Research Comment on “Global Surgery - Informing National Strategies for Scaling Up Surgery in Sub-Saharan Africa.” Int J Health Policy Manag. 2019;8: 58–60. doi:10.15171/ijhpm.2018.106

13. Ross O, Shakya R, Shrestha R, Shah S, Pradhan A, Shrestha R, et al. Pathways to effective surgical coverage in a lower-middle-income country: A multiple methods study of the family physician-led generalist surgical team in rural Nepal. PLOS Glob Public Health. 2023;3: e0001510. doi:10.1371/journal.pgph.0001510

14. Kumar N, Hyman GY, Gerk A, Wurdeman T, Uribe-Leitz T, Corlew S, et al. Scalable policy adoption and sustainable implementation of surgical care in the Americas. Rev Panam Salud Publica. 2024;48: e132. doi:10.26633/RPSP.2024.132

15. Willer BL, Mpody C, Tobias JD, Nafiu OO. Association of Race and Family Socioeconomic Status With Pediatric Postoperative Mortality. JAMA Netw Open. 2022;5: e222989. doi:10.1001/jamanetworkopen.2022.2989

16. Bouchard ME, Kan K, Tian Y, Casale M, Smith T, De Boer C, et al. Association Between Neighborhood-Level Social Determinants of Health and Access to Pediatric Appendicitis Care. JAMA Netw Open. 2022;5: e2148865. doi:10.1001/jamanetworkopen.2021.48865

17. Haft M, Schmerler J, Prieskorn BP, Murdock CJ, Nelson S, Srikumaran U, et al. Surgical Disparities in the United States from 2012 to 2020: Race and Payer Status are Associated with Increased Time to Urgent and Emergent Surgery. J Racial Ethn Health Disparities. 2025. doi:10.1007/s40615-025-02489-4

18. Michelson KA, Bachur RG, Rangel SJ, Finkelstein JA, Monuteaux MC, Goyal MK. Disparities in Diagnostic Timeliness and Outcomes of Pediatric Appendicitis. JAMA Netw Open. 2024;7: e2353667. doi:10.1001/jamanetworkopen.2023.53667

